# Enhancing deep sleep with auditory stimulation to boost cognitive training responsiveness in people with Parkinson’s disease and mild cognitive impairment: Rationale and design of the double-blind randomized controlled trial PD-CogT-Sleep

**DOI:** 10.64898/2026.11.02.26367082

**Authors:** Anja Ophey, Marie Therese Kleinsorge, Noemi A. Wallimann, Angelina Maric, Sandra V. Loosli, Brit Mollenhauer, Bettina Balint, Reto Huber, Christian R. Baumann, Elke Kalbe, Simon J. Schreiner

## Abstract

**Background:** Cognitive training (CogT) is an established non-pharmacological intervention for people with Parkinson’s disease (PD) and mild cognitive impairment (PD-MCI). However, overall effects on cognitive outcomes are small to moderate and interindividually highly heterogeneous. Sleep –especially deep sleep–could be an important determinant of CogT responsiveness in PD, and may have beneficial effects on PD pathophysiology. Therefore, the enhancement of deep sleep by phase-targeted auditory stimulation (PTAS) could strengthen the effects of CogT on cognitive symptoms and counteract disease progression.

**Methods:** PD-CogT-Sleep is a monocentric, partly decentralized, double-blind, randomized sham-controlled trial that combines two remotely delivered non-pharmacological interventions, to assess the additive effect of PTAS on digital CogT responsiveness in people with PD-MCI. The primary outcome is executive functions; Secondary and exploratory outcomes include further cognitive functions, participant-reported outcome measures, blood-based biomarkers, and high-density sleep electroencephalography markers. Outcomes are assessed at baseline, five weeks post-allocation and at 3-month follow-up. We aim to consecutively recruit *N*=50 participants, who will be randomized 1:1 to receive CogT+PTAS or CogT+Sham.

**Discussion:** The PD-CogT-Sleep study integrates wearable and digital technologies to systematically combine a novel, non-invasive, sleep-enhancing intervention (PTAS) with established CogT to improve cognition and evaluate PTAS’ disease-modifying potential through its effects on blood-based biomarkers in PD-MCI. The expected results will foster our understanding of the interplay of sleep, cognition, and neurodegeneration, and may guide further development of safe, feasible, scalable, patient-centered, technology-based non-pharmacological interventions counteracting cognitive decline in neurodegenerative diseases.

**GRAPHICAL ABSTRACT:** 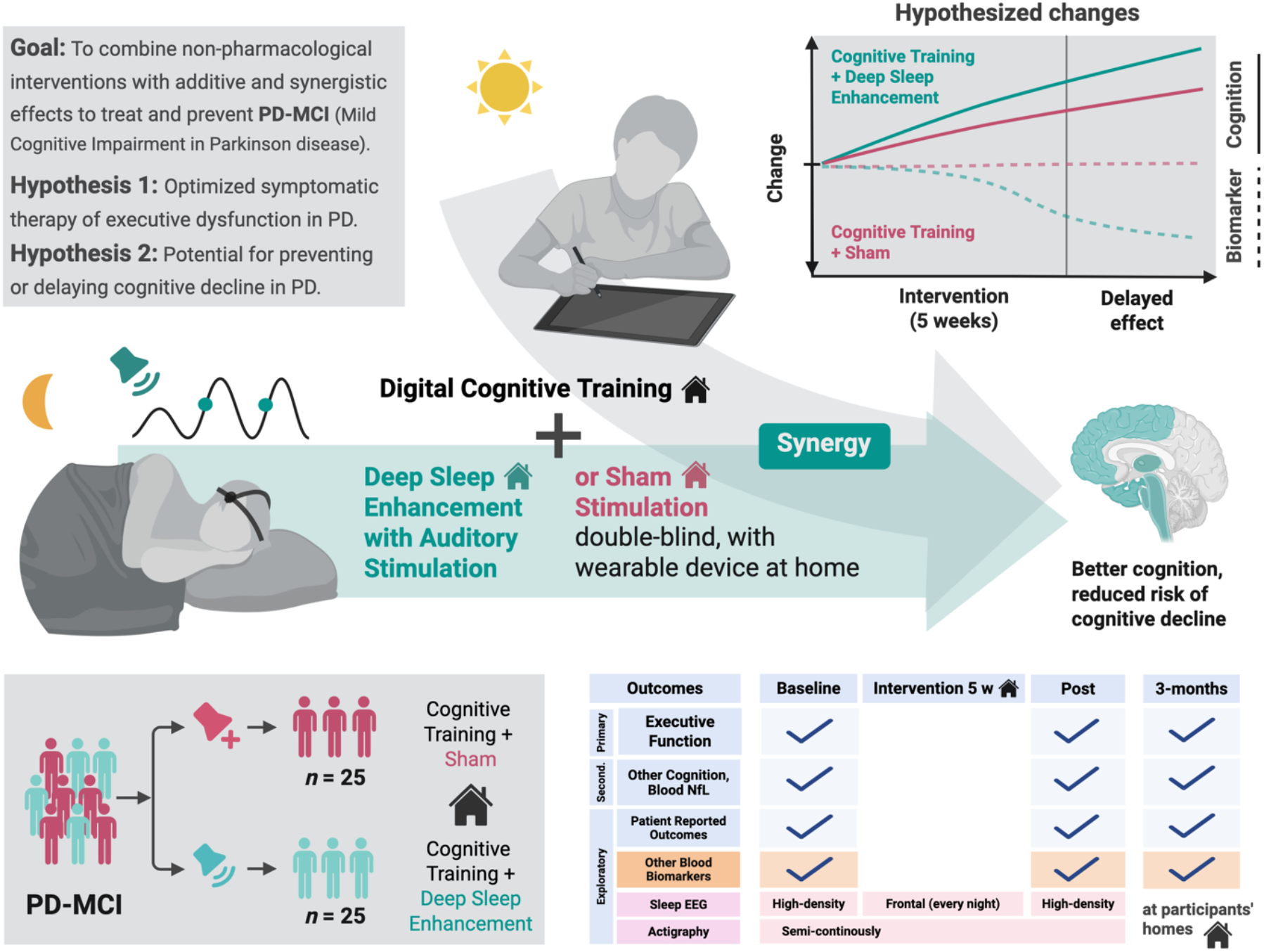

Study Design Overview: PD-CogT-Sleep is a randomized controlled double-blind clinical trial on the effects of deep sleep enhancement with phase-targeted auditory stimulation vs. sham on the responsiveness of cognitive training. Created in https://BioRender.com

**HIGHLIGHTS:**

- Effects of cognitive training (CogT) in Parkinson’s disease (PD) are heterogeneous
- Deep sleep could be an important determinant of CogT responsiveness, but is altered in PD
- Phase-targeted auditory stimulation (PTAS) enhances slow waves to restore deep sleep
- Combining wearable and digital technology allows remote PTAS and CogT application
- PD-CogT-Sleep is a double-blind, partly decentralized RCT on CogT+PTAS vs. CogT+Sham

## BACKGROUND

Cognitive decline is a common and debilitating non-motor symptom for people living with Parkinson’s disease (PD), the second most frequent neurodegenerative disorder.^1,2^ Cross-sectionally, mild cognitive impairment (MCI) is present in about 40% of people living with PD (PD-MCI),^3^ and longitudinally up to 80% develop PD dementia (PDD).^4,5^ PD-related cognitive impairment is thought to result from overlapping neurodegenerative processes, including cortical Lewy body pathology, Alzheimer’s co-pathology, vascular injury, and neurotransmitter dysfunction.^6^ There is increasing evidence that blood-based biomarkers such as neurofilament light chain (NfL), a marker of axonal damage, have prognostic value, as they predict and correlate with cognitive decline in PD.^7–9^ To date, pharmacotherapy against cognitive decline in PD is limited and not recommended or approved for PD-MCI, although the latter are in need of therapies and secondary prevention.^2,10^

Non-pharmacological interventions including cognitive training (CogT) are increasingly recognized as therapeutic options counteracting cognitive decline.^10–13^ In this regard, digital CogT offers great potential due to its flexibility, adaptability, accessibility, and scalability.^12,14^ Meta-analyses demonstrated positive effects of CogT on cognitive functions in people living with PD,^11,13^ with predominant effects on executive and other cognitive domains vulnerable in PD. Overall, however, the behavioral effects of CogT are modest and highly heterogeneous.^15^ While some predictors of CogT responsiveness, such as lower baseline performance and better overall cognition,^16^ have been identified, a substantial proportion of variance remains unexplained.^17,18^ Understanding determinants and predictors of responsiveness to CogT is of high clinical importance to guide precision medicine approaches in vulnerable target groups, such as PD.

Sleep, especially deep sleep, could be an important determinant of CogT responsiveness. Deep sleep supports learning and cognitive functioning,^19^ and accumulating evidence suggests a reciprocal relationship between CogT and deep sleep: Habitual sleep between training sessions is required for successful working memory training, indicating that sleep consolidates training-induced cognitive processes.^20^ Likewise, CogT induces local increases in slow-wave activity (SWA), a marker of sleep depth and plasticity measured with cortical surface electroencephalography (EEG), which predict CogT responsiveness.^21^ In people living with PD, deeper night-time sleep between days of training sessions is associated with greater responsiveness to working memory training.^22^ Together, these findings suggests that deep sleep may enhance, while poor sleep may hinder responsiveness to CogT, especially in people living with PD.

A second line of evidence suggests that deep sleep may play a mechanistic role in neurodegenerative diseases by promoting processes that counteract neurodegeneration.^23^ Sleep regulates levels of critical proteins implicated in PD and other neurodegenerative diseases, possibly by promoting their clearance or reducing their production.^24–26^ In people living with PD, deep sleep is impaired, particularly in those with cognitive impairment,^27–29^ and has been linked to excessive daytime sleepiness, dyskinesia, and faster disease progression.^30–36^ Deep sleep is defined by the occurrence of slow waves – low-frequency (0.5-4.5 Hz), large-amplitude (>75 μV) EEG oscillations reflecting synchronized thalamocortical and cortical neuronal activity.^37^ Their abundance and amplitudes are intimately linked to the beneficial effects of deep sleep.^37,38^ However, slow waves diminish with aging and neurodegeneration and may both reflect and contribute to brain changes and cognitive decline.^39^ Intriguingly, pharmacologically enhancing deep sleep reduced α-synuclein and ß-amyloid pathology in mice^40,41^ and improved sleep-wake disturbances in people living with PD.^42–44^ Overall, deep sleep may contribute to symptom burden in PD and deep sleep enhancement represents a promising therapeutic strategy for targeting cognitive decline and potentially modifying disease progression.^19,23,38,45,46^ In this context, non-pharmacological interventions offer potential advantages in safety and long-term applicability over pharmacological interventions.

Phase-targeted auditory stimulation (PTAS) of sleep slow waves is a novel non-pharmacological method for enhancing deep sleep.^47^ PTAS delivers non-arousing tones to the up-phase of endogenous slow waves, thereby increasing slow-wave amplitude, measurable by increased SWA. Intriguingly, PTAS also enhances deep sleep-related processes, such as positive dynamics of ß-amyloid,^48^ executive performance on the next day,^49^ or overnight memory retention,^47,50^ even in older adults and individuals with MCI.^51,52^ Compared with other slow-wave enhancement approaches, PTAS has no known side effects, no or only minimal effects on sleep macroarchitecture, permits simultaneous EEG recording without stimulation artifacts, enables blinded sham-controlled designs, and can be applied unobtrusively across multiple nights.^45,53–55^ Wearable devices have further enabled safe and effective multi-night, home-based PTAS in older adults and people living with Alzheimer’s disease or PD.^42,44,56–58^ Therefore, wearable-based PTAS is a promising strategy for long-term, non-invasive, non-pharmacological enhancement of deep sleep in aging and neurodegenerative disease.

To summarize, therapeutic enhancement of deep sleep could strengthen the effects of CogT for people living with PD, improve cognitive symptoms, and may enable disease-modification as indicated through its potential effects on blood-based biomarkers. The PD-CogT-Sleep study aims to assess the effects of deep sleep and its enhancement by PTAS on CogT responsiveness and blood-based biomarkers related to cognitive decline in people living with PD-MCI. This monocentric, partly decentralized, double-blind, randomized controlled trial combines digital, home-based, adaptive, multi-domain CogT with home-based PTAS or sham stimulation using wearable devices in a parallel group design. The following main research questions will be addressed:

**1. Does deep sleep enhancement with PTAS improve CogT responsiveness in people with PD-MCI?**
**2. Does deep sleep enhancement with PTAS in addition to CogT induce changes in blood-based biomarkers such as NfL, potentially indicating a reduced risk of further cognitive decline?**

Reporting of this study protocol follows the “Standard Protocol Items: Recommendations for Interventional Trials” (SPIRIT) checklist.^59^

## METHODS

### Study Design and Setting

The PD-CogT-Sleep study is designed as a monocentric, partly decentralized, double-blind, parallel two-arm, randomized controlled trial assessing the additive effects of digital, home-based, adaptive, multi-domain CogT and home-based PTAS using wearable devices (CogT+PTAS) versus sham (CogT+Sham) on clinical and biomarker outcomes (superiority design) in people living with PD-MCI five weeks post-allocation (t1) and at 3-month follow-up (t2). We aim to consecutively recruit *N*=50 participants that will be randomized in a 1:1 ratio to the CogT+PTAS vs. CogT+Sham group. The sham-controlled design allows for blinding of both participants and outcome assessors, i.e., the study will be conducted under double-blind conditions. Inclusion and exclusion criteria will be assessed during a screening visit (t-1) and the screening adaptation nights with the PTAS device at the participants’ homes. Standardized, validated assessments and questionnaires will be used to assess primary, secondary, and exploratory outcomes at baseline (t0), t1, and t2. The combination of two remotely delivered interventions (digital, home-based, adaptive, multi-domain CogT and home-based PTAS) offers accessible and scalable non-pharmacological approaches targeting cognitive symptoms of PD.

Figure 1 visualizes the planned study design including study procedures, that are further outlined in the following chapters.

**Figure 1.**
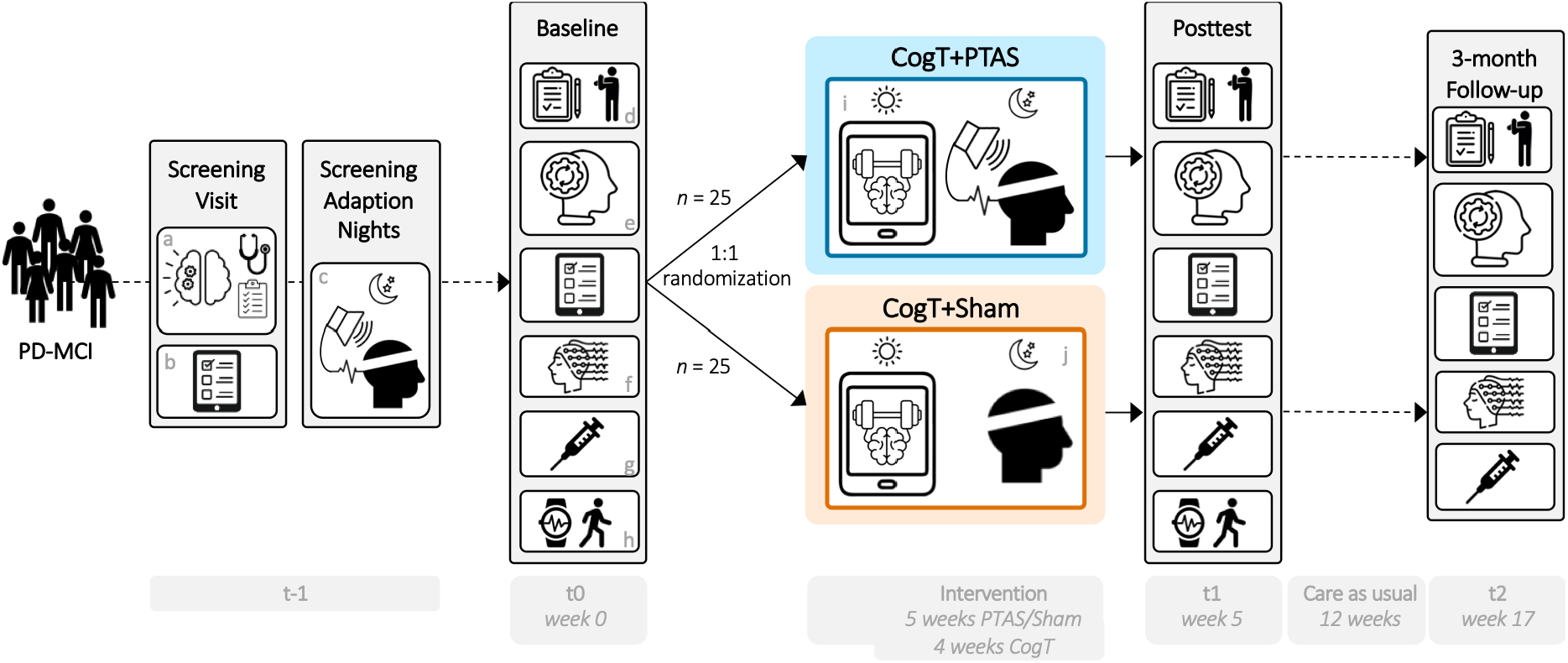
Study Design of the PD-CogT-Sleep Study. CogT, cognitive training; PD-MCI, Parkinson’s disease with mild cognitive impairment; PTAS, phase-targeted auditory stimulation. a. demographic and general clinical assessments during screening b. questionnaires / participant reported outcome measures (PROMs) c. PTAS and multi-night frontal sleep EEG d. update medical history e. cognitive assessment f. sleep high-density electroencephalography (hdEEG) g. blood sampling h. actigraphy i. digital cognitive training j. sham stimulation and multi-night frontal sleep EEG

### Ethics and Trial Status

Ethical approval was granted by the local ethics committee (Kantonale Ethikkommission Zurich, Switzerland) on 2025-12-18 (Identifier: 2025-D0081). Additionally, the study was approved by Swissmedic, the Swiss authority responsible for the authorization and supervision of therapeutic products, on 2025-12-05 (Identifier: 10001523). Simon J. Schreiner, University Hospital Zurich, is the sponsor-investigator. Written informed consent to participate will be obtained from all participants. The study was prospectively registered on clinicaltrials.gov (Identifier: NCT07441915) on 2026-12-02. The protocol version is V2.3 as of 2026-04-15.

The trial status is ongoing with the first participant included on 2026-04-22. We expect the recruitment to be finished by December 2027.

### Recruitment and Informed Consent

Participants are recruited via the outpatient clinic at the University Hospital Zurich, Switzerland, advertisements in patient organization newsletters across Switzerland, and referrals from clinicians. A study flyer serves as a first comprehensive information sheet outlining the details of PD-CogT-Sleep, and, in the case of interest in study participation, further details are discussed via telephone or in face-to-face contact following the clinical visits in the outpatient clinic. All relevant aspects of the study –its nature, purpose, procedures, risks, benefits, and voluntary participation–will be clearly explained to potential participants. Adequate time for consideration and consultation will be provided. Informed consent will be obtained prior to any study-specific procedures.

### Inclusion and Exclusion Criteria

We aim to include *N*=50 people living with PD-MCI defined by level I criteria of the Movement Disorder Society (MDS) for PD-MCI,^60^ namely reported or observed gradual cognitive decline and objective impairment on cognitive screening. The main inclusion criteria are (i) clinical diagnosis of PD along MDS criteria^1^; (ii) PD-MCI according to the MDS criteria (level I) for PD-MCI^60^: Montréal Cognitive Assessment (MoCA)^61^ 18-26 points^62^; (iii) sufficient German language comprehension; (iv) stable dosing of dopaminergic and other PD treatment ≥ 14 days prior to the intervention period. The main exclusion criteria are (i) a clinical diagnosis of dementia; (ii) known presence of relevant neurologic (other than PD) or psychiatric disorders; (iii) regular use of benzodiazepines and other central nervous system (CNS)-depressant substances; (iv) sleep disorders that could interfere with PTAS; (v) absent EEG engagement to PTAS during screening nights. For a complete list of inclusion and exclusion criteria, please refer to the Supplementary Material 1.

### Randomization and Blinding

The randomization will be based on a minimization procedure designed to minimize differences between the CogT+PTAS and CogT+Sham group in age, sex, and global cognition as assessed by the MoCA.^61^ The minimization randomization is implemented using the R package *Minirand.*^63^ We aim at a 1:1 ratio of the number of subjects in the CogT+PTAS and CogT+Sham group. The covariates age, sex, and MoCA total score will be equally weighted (1/3 each). The participants will be consecutively randomized following the baseline assessment by a person not involved in data collection.

PD-CogT-Sleep is a double-blind trial. The allocation sequence is concealed through automated randomization logic, with group assignment communicated only to the technical staff responsible for the PTAS device configuration. Study physicians and study personnel assessing outcomes will not have access to the randomization allocation list or to the documentation of the programmed PTAS equipment. The technical staff members and their supervisors, who are unblinded with regard to the study arm, will not perform any outcome assessments. Any violations of blinding will be documented.

### Interventions

The PD-CogT-Sleep intervention period includes 5 weeks of decentralized interventions, including 5 weeks of home-based PTAS or sham stimulation using wearable devices and 4 weeks of digital, home-based, adaptive, multi-domain CogT starting from the second week. This stepwise increase in intervention complexity will contribute to the overall feasibility of the combined intervention for individuals with PD-MCI. Additionally, this design allows to explore CogT-related changes in frontal sleep EEG features in the CogT+Sham group at high temporal resolution (exploratory outcome). Throughout the clinical investigation, participants may continue to receive standard medical care for PD and other comorbid conditions, provided that these treatments remain stable and do not interfere with the primary or secondary outcomes related to sleep and cognition. All concomitant treatments will be recorded, including medication name, dosage, frequency, indication, and any changes made during the course of the study.

#### Slow Wave Enhancement through PTAS or Sham Stimulation

Participants will be instructed to use the latest version of the wearable device TOSOO Axora by TOSOO AG (Zurich, Switzerland, https://www.tosoo.ch) during their regular sleep times every night at their homes starting in the first week of the intervention. The TOSOO Axora is a wearable headband for automated application of PTAS based on continuous EEG recording, NREM sleep detection, and estimation of ongoing slow waves.^64^ For PTAS, 50-millisecond bursts of pink noise (∼50 dB, maximum 80 dB) are delivered synchronised to the up-phase of detected slow waves to enhance slow waves. For sham stimulation, tones are delivered in a way that does not enhance slow waves, maintaining participant and assessor blinded, while permitting standardized longitudinal EEG collection. To preserve blinding in case participants inadvertently perceive a tone, the exact sham stimulation procedure is only known to unblinded study personnel. All participants will wear the headband nightly for 5 weeks. The PTAS vs. sham configuration will start from the beginning of the intervention.

Configurable thresholds allow tailoring the application of PTAS to altered sleep characteristics, such as low-amplitude slow waves, in aging and neurological conditions, including PD.^44,56,65^ Algorithm thresholds and volume settings are personalized during the screening adaptation nights further described below to achieve neurophysiological target engagement, defined as a significant auditory evoked potential between ON and OFF windows (intervals during which stimulation is delivered vs. omitted within the same night) in at least one adaptation night. Adherence to the use of the TOSOO Axora device is automatically tracked through the device’s internal logging system, which assesses nightly wear time, stimulation events, and EEG signal quality.

Before the main intervention period, up to eight screening adaptation nights following a successful screening visit and enrolment serve two purposes: (i) training participants in the use of the TOSOO Axora device and (ii) optimizing sleep detection and stimulation settings. Participants will be retrained during the intervention period if difficulties with device handling arise. Data from the screening nights will be analyzed to refine and personalize stimulation parameters as to maximize precision. The device continuously monitors for signs of arousal and dynamically adjusts sound volume to minimize disruptions. The number of screening nights (up to eight, typically three to five) will depend on the extent of required optimization.

#### Digital Cognitive Training

All participants will receive digital, home-based, adaptive, multi-domain CogT, delivered by the CE-certified Class I medical device HeadApp/NEUROvitalis (HelferApp GmbH, Gommern, Germany https://start-headapp.helferservices.net), which is accessible at home via tablet with internet access. During the t0 study visit, participants will get pseudonymized access-data for the HeadApp, which will be pre-installed on the tablets participants receive for the duration of the decentralized intervention.

The CogT application has recently been evaluated in both prodromal and advanced phases of PD.^66,67^ The total CogT duration will be 600 minutes (3×50 min/week over four weeks), comparable to previous protocols and recommendations.^15,66–68^ Participants will be instructed to train 12 sessions over four weeks (i.e., 3 sessions per week), starting in the second week of the intervention time period and to evenly distribute the training sessions across time while maintaining flexibility on choosing individual training days and specific training times. CogT adherence will be monitored weekly by the study technician on the HeadApp website and any queries of participants will be answered by telephone or email, depending on the participant’s choice.

The CogT is designed as a multi-domain CogT that continuously adapts its difficulty and complexity to the individual cognitive profile of the participant. The training tasks are based on the scientifically validated NEUROvitalis program (ProLog, Therapie und Lernmittel GmbH, Cologne).^69–73^ During each training session, five out of six cognitive tasks from HeadApp/NEUROvitalis will be trained for 10 minutes each. The task selection will follow a pre-specified training plan across participants, ensuring that each participant trains with the same tasks. The level of difficulty in each task, however, adapts to the individual performance to optimally balance engagement while avoiding frustration. The final level of task difficulty in one session will be the starting level of difficulty in the following training session. With increasing levels of difficulty, all training tasks involve an increasing focus on executive functions, constituting the primary endpoint of our study.

#### Non-Compliance

Non-compliance with the sleep device application is defined as applying the device during (i) less than 25 of 35 total nights (i.e., >30% of missed nights over 5 weeks) or (ii) less than 5 of 7 nights in week 5, i.e., in the final week before the follow-up visit. Non-compliance with the CogT is defined as completing (i) less than 10 of 12 total sessions or (ii) less than 480 of 600 total minutes (i.e., >20% missed CogT) of planned CogT over 4 weeks. Furthermore, non-compliance indicated by intentional or repeated incorrect use of the device or training platform despite guidance may lead to exclusion from per-protocol analyses, but participants will remain in safety follow-up unless withdrawn.

### Outcomes and Assessments

The primary outcome is the change in executive performance (i.e., an equally weighted composite domain score) between t0 and follow-up timepoints t1 and t2 compared between groups (CogT+PTAS vs. CogT+Sham). Secondary outcomes are corresponding changes in global cognition (equally weighted composite score across cognitive domains), other cognitive functions (equally weighted composite scores of attention & working memory, memory, visuo-cognition, language), and the blood-based biomarker NfL. Exploratory endpoints include relationships among clinical, sleep, and blood biomarker features, and change between t0 and follow-up timepoints t1 and t2 compared between groups (CogT+PTAS vs. CogT+Sham) in subjective cognitive decline (SCD), quality of life, non-motor outcomes (i.e., fatigue, depressive symptoms, sleep disturbances, daytime sleepiness), vigilance, inhibitory control, blood-based biomarkers other than NfL to track other neurodegenerative features (e.g., α-synuclein, ß-amyloid40/42, tau, phospho-tau, neuroinflammatory markers), frontal sleep EEG markers (i.e., sleep parameters derived from the TOSOO Axora device worn during the intervention), local sleep EEG markers, such as local increases in SWA (derived from high-density sleep EEG (hdEEG) recordings during study visits), physical activity and circadian rhythm (derived from actigraphy). A full list of assessments conducted in the PD-CogT-Sleep study can be assessed in Supplementary Material 2.

### Safety Aspects

All device deficiencies, serious adverse events (SAEs), and non-serious adverse events (AEs) will be documented and fully investigated throughout the entire study period, from informed consent until the final study procedure. Both home-based PTAS using wearable devices and digital, home-based, adaptive, multi-domain CogT are non-invasive interventions associated with no or minimal risk, and all study assessments represent routine low-risk procedures. Risk management and safety monitoring will be conducted in accordance with Good Clinical Practice and applicable regulatory requirements.

### Sample Size and Power Calculation

To determine an adequate sample size, an a-priori power analysis was conducted with G*Power^74^ (http://www.gpower.hhu.de). The study will be powered to reliably detect behavioral effects in the domain of executive functions (the primary outcome). We a priori assumed a small-to-medium effect size in the primary outcome compared between the CogT+PTAS and the CogT+Sham. A repeated-measures ANOVA (within–between interaction; *f*=0.175; 3 timepoints, α=0.05; 80% power), considering the strong correlation between two repeated measures of executive functions (r=0.6)^75^, indicated a required sample size of 44 participants. Accounting for an anticipated dropout rate of 14%, comparable to previous intervention studies, the final target sample size was set at 50 participants.

### Data Management and Statistical Analysis

Regular monitoring visits will be performed by the Clinical Trials Center (CTC) of the University Hospital Zurich, Switzerland. All patient information and study data are to be handled strictly confidentially and managed in accordance with Good Clinical Practice and applicable data protection regulations. All study data will be collected using validated electronic case report forms in a REDCap and MyCap environment with pseudonymization, role-based access control, and complete audit trails. Source data from the wearable sleep device, hdEEG, actigraphy, and blood samples will be handled, transferred, and stored securely under the same regulatory framework.

We will analyze data of all participants who complete at least one follow-up (t1, t2) in an intention-to-treat (ITT) analysis. Effects on primary, secondary and exploratory outcomes will be assessed with linear mixed models (LMM) including time (t0, t1, t2), group (CogT+PTAS vs CogT+Sham), and the interaction between time and group as fixed factors and the respective outcome measure as dependent variable. We will perform model diagnostics and in case we see clear deviations from normality or homoscedasticity, we will perform a robust version of the linear mixed model analysis and non-parametric post-hoc testing. P-values lower than 0.05 are considered significant, p<0.1 and p≥0.05 are considered trend-level. Exploratory subgroup analyses will evaluate whether participants’ baseline characteristics (e.g., demographic characteristics such as age and sex, clinical characteristics such as PD stage and subtype, cognitive performance) have an effect on responsiveness to CogT+PTAS vs. CogT+Sham by including these variables as additional co-variates in the LMM. Any deviation(s) from the original statistical analysis plan will be clearly documented and reported in the final clinical investigation report if they occur during or after data analysis. We will conduct a complete-case analysis. Drop-outs were considered in the a priori sample size calculation and will not be replaced. We may still be analyzing data from participants in whom singular data from exploratory outcomes (e.g., questionnaires) are missing.

## DISCUSSION

The PD-CogT-Sleep study combines deep-sleep enhancement by home-based PTAS with established digital, home-based, adaptive, multi-domain CogT in people with PD-MCI, who are at high risk of dementia. PTAS is a novel, non-invasive intervention for enhancing deep sleep that can be unobtrusively delivered at the participants’ homes using wearable devices. Thereby, this partially decentralized randomized controlled clinical trial tests whether combining CogT with deep-sleep enhancement by PTAS strengthens the effect of CogT on cognitive functions, and evaluates the potential of PTAS for disease-modification and slowing of cognitive decline as indicated by changes in blood-based biomarker levels, such as NfL.

The expected results will provide important insights of relevance for future clinical trials, clinical neuroscience, and the pathophysiology and alleviation of PD-MCI: First, PD-CogT-Sleep could inspire the development of technology-based non-pharmacological strategies for the treatment and management of PD-MCI. These strategies will be of relevance for other conditions with concurrent impairment of deep sleep and cognition, e.g., Alzheimer’s disease or Attention Deficit Hyperactivity Disorder. Second, the blood-based biomarker outcomes will provide novel insights into the neurobiological effects of PTAS in the setting of PD-MCI and CogT. Modifications of blood-based biomarkers, such as NfL –a marker of axonal damage and future cognitive decline in PD^7–9^–, would indicate the potential of PTAS for prevention of dementia and disease-modification.

The study was designed to rigorously assess the additive effect of PTAS on CogT responsiveness in individuals with PD-MCI. We chose this combination of interventions because CogT and PTAS may not only provide additive but also synergistic effects on cognitive outcomes: Enhancing deep sleep with PTAS may strengthen the effects of CogT by improving offline consolidation during sleep and vigilance during training. In turn, CogT could increase SWA through use-dependent mechanisms, which may further strengthen the effects of PTAS.^21,56,76^ The use of home-based PTAS using wearable devices is grounded in previous research demonstrating its feasibility, safety, and potential cognitive benefits in older adults and patient populations, including PD.^44,56,57^ The wearable format allows for consistent, home-based use without medical supervision, which supports ecological validity and scalability.^55^ Combining two decentralized interventions (digital CogT and home-based PTAS) therefore evaluates two promising, scalable, non-pharmacological strategies for secondary dementia prevention in PD. Given the overall demanding study protocol, the trial will also provide important insights into the feasibility of combined remote interventions in vulnerable clinical populations.

The parallel-group design avoids carryover effects and is suited for interventions with long-lasting or training-based outcomes. The sham control condition (identical device use at night without PTAS) ensures blinding and controls for placebo effects. The double-blind setup minimizes bias from both participants and assessors.

To summarize, investigating the modulating effect of deep sleep enhancement by home-based PTAS on CogT responsiveness will contribute to our understanding of the interplay of sleep, cognition, and neurodegeneration under real-world conditions. Through its unique study design, the PD-CogT-Sleep study may support the development of novel patient-centered, technology-based non-pharmacological interventions targeting unmet needs in neurodegenerative disease.

## Data Availability

This is a study protocol and no data was published yet.

## ACKNOWLEDGEMENTS

We thank the Jacques and Gloria Gossweiler Foundation (main sponsor) and Theodor and Ida Herzog-Egli Foundation for the financial support of the PD-CogT-Sleep study. The PD-CogT-Sleep study is conducted as a joint cooperation between University Hospital Zurich, University of Zurich, and TOSOO AG. We thank the entire team of the TOSOO AG, the Clinical Trials Center (CTC), and regulatory authorities for their valuable support.

## AUTHOR CONTRIBUTIONS

AO: Conceptualization, Funding acquisition, Methodology, Project administration, Investigation, Writing – original draft

MTK: Methodology, Project administration, Investigation, Writing – review and editing

NAW: Project administration, Investigation, Writing – review and editing

AM: Methodology, Software, Writing – review and editing

RH: Methodology, Software, Supervision, Writing – review and editing

SL: Methodology, Resources, Supervision, Writing – review and editing

BM: Methodology, Resources, Supervision, Writing – review and editing

BB: Conceptualization, Resources, Writing – review and editing

CB: Conceptualization, Resources, Supervision, Writing – review and editing

EK: Conceptualization, Resources, Supervision, Writing – review and editing

SJS: Conceptualization, Funding acquisition, Methodology, Project administration, Resources, Supervision, Writing – original draft

