## Supplementary for "Enhancing deep sleep with auditory stimulation to boost cognitive training responsiveness in people with Parkinson’s disease and mild cognitive impairment: Rationale and design of the double-blind randomized controlled trial PD-CogT-Sleep"

---

**Supplementary Material 1.** Full List of Inclusion and Exclusion Criteria ------------ p. 2

**Supplementary Material 2.** Assessments in the PD-CogT-Sleep Study ------------ p. 4

**Supplementary References** ---------------------------------------------------------------- p. 8

---

### *Supplementary Material 1.* **Full List of Inclusion and Exclusion Criteria**

The following inclusion criteria will be applied:

- **clinical diagnosis of Parkinson's disease (PD)** along the Movement Disorder Society (MDS) criteria [1]
- **mild cognitive impairment (MCI) associated with PD (PD-MCI) according to the MDS criteria (level I)**[2]: Montréal Cognitive Assessment (MoCA) 18-26[3]
- **stable home situation** (e.g. long-term place to live) that allows for reliable application of intervention for the duration of the study
- **ability to apply the sleep intervention** for the duration of study, either alone or with assistance of a co-habitant if needed
- **ability to apply the cognitive training (CogT) intervention** for the duration of study
- **sufficient German language comprehension** to follow the study procedures and answer all questions related to the study outcomes
- **stable dosing of dopaminergic and other PD treatment** ≥ 14 days prior to the intervention period, which will be expected to remain stable until the end of the study

The following exclusion criteria will be applied:

- **Diagnosis/Comorbidities:**
    - clinical diagnosis of dementia (cognitive impairment sufficient to interfere with independence in everyday activities, i.e. "major neurocognitive disorder", DSM-5)
    - known presence of neurologic (other than PD) or psychiatric disorder
    - Parkinsonism without response to levodopa or atypical Parkinsonian syndromes as assessed from medical history and clinical examination
    - severe medical conditions (for example, renal insufficiency, liver failure, or congestive heart failure) as assessed in the semi-structured screening interview
    - regular use of benzodiazepines and other central nervous system (CNS)-depressant substances as assessed in the semi-structured screening interview
    - known or suspected drug- or medication abuse as assessed in the semi-structured screening interview
    - substance or alcohol abuse (i.e. > 0.5 l wine or 1 l beer per day) as assessed in the semi-structured screening interview
- **Sleep disorders that could interfere with the sleep intervention:**
    - obstructive sleep apnea with apnea-hypopnea index (AHI) >15, apnea-related NREM sleep fragmentation, and indication for treatment (in turn, primarily REM-related sleep apnea, not requiring specific treatment may be considered eligible); or use of continuous positive airway pressure (CPAP)
    - Restless Legs Syndrome
    - frequent (i.e. weekly) non-REM sleep parasomnia

- **Phase-targeted auditory stimulation (PTAS):**
  - inability to hear the tones produced by the sleep headband (TOSOO Axora device)
  - neurophysiological target engagement, defined as a significant auditory evoked potential between ON and OFF windows (intervals during which stimulation is delivered vs. omitted within the same night) in at least one screening adaptation night
  - skin disorders/problems/allergies/sensitivities, including sensitivities to acrylates, silver/silver chloride or liquid wet gel in general, in face/ear area that could worsen with electrode application
  - known or suspected non-compliance

- **Cognition & informed consent:**
  - inability to follow the procedures of the study, e.g. due to language problems, cognitive deficits
  - failure to give informed consent

- **Other studies:**
  - participation in another study with investigational interventions within 30 days (PTAS) or 1 year (CogT) preceding and during the present study
  - previous enrolment in the current study
  - enrolment of the investigator, his/her family members, employees and other dependent persons

- **Special events / behavior with impact on circadian rhythm, sleep, or cognition:**
  - shift work (work during the night)
  - travelling more than 2 time zones in the last month before intervention starts or during intervention (start of intervention will be adapted to fit with this criteria)
  - planned medical intervention of substantial relevance, e.g. surgery, during intervention (routine assessments, e.g. check-ups will be allowed)

- **Pregnancy:**
  - women who are pregnant or breastfeeding,
  - intention to become pregnant during the course of the study,
  - lack of safe contraception, defined as: Female participants of childbearing potential, not using and not willing to continue using a medically reliable method of contraception for the entire study duration, such as oral, injectable, or implantable contraceptives, or intrauterine contraceptive devices, or who are not using any other method considered sufficiently reliable by the investigator in individual cases. Please note that female participants who are surgically sterilized/hysterectomized or post-menopausal for longer than 1 year are not considered as being of child bearing potential.

*Supplementary Material 2.* **Assessments in the PD-CogT-Sleep**

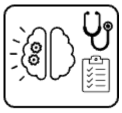
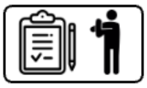

**Demographic and general clinical assessments during screening.** We will obtain the following information during screening using a semi-structured interview and questionnaires:

- demographic information (age, sex, education, living situation)
- inclusion/exclusion criteria
- more detailed information about health status (e.g., current medication, medical history, daily habits, chronotype, and handedness)
- PD-specific details (disease duration, dominant body side, current motor [MDS-UPDRS-III[4]] and medication status)
- cognitive status with level I cognitive assessment: MoCA [5]
- PD-related non-motor symptoms: Non-motor Symptom Questionnaire (NMSQ)[6]
- PD-related sleep disturbances: Parkinson's Disease Sleep Scale 2 (PDSS-2)[7]
- sleep quality: Pittsburgh Sleep Quality Index (PSQI)[8]
- daytime sleepiness: Epworth Sleepiness Scale (ESS)[9]
- cognitive reserve: Cognitive Reserve Index questionnaire (CRIq)[10]

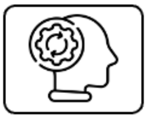

**Cognitive assessment.** The cognitive test battery will be applied at t0, t1 and t2 during the clinical visit by study personnel (e.g., study physician, students of Medicine or Psychology) trained and supervised by a neuropsychologist. The whole test battery will take 90 minutes, including a 10-minute break. The test battery allows for a level II cognitive assessment of PD-MCI following the established MDS criteria for PD-MCI,[2] adapted according to recommendations for cognitive assessments for people with PD in German speaking countries.[11,12] Table 1 lists the analyzed test scores grouped to their respective cognitive domain including an indication, if parallel versions, alternating between time points, will be used. Published, established norms accounting for demographic influences (e.g., age, sex, education) will be applied to raw scores of the cognitive test battery. All statistical evaluations with cognitive outcomes will then be based on these standardized values transformed to z-scores for a unified scale range across assessments. To build the cognitive composite scores, the single demographically adjusted, standardized z-scores will be summed up and divided by the number of tests in the respective domain. In addition to the described test battery, we will assess the participants' current vigilance state with additional tests on a tablet:

- momentary sleepiness: Karolinska Sleepiness Scale (KSS)[13,14]
- vigilance: Sustained Attention to Response Task (SART)[15] and Psychomotor Vigilance Task (PVT)[16]

*Table 1.* Cognitive test battery in PD-CogT-Sleep

| Domain / Subdomain | Test Abbre-viation | Full test name | Normative Data / Test Reference | Parallel versions |
|---|---|---|---|---|
| **Executive Functions** | | | | |
| Semantic verbal fluency | RWT sem | Regensburger Wortflüssigkeitstest | Aschenbrenner, Tucha and Lange [17] | X Food or Animals |
| Phonemic verbal fluency | RWT phon | Regensburger Wortflüssigkeitstest | Aschenbrenner, Tucha and Lange [17] | X P- or S-words |
| Set-Shifting | TMTB/A | CERAD Trail Making Test (TMT): TMT-B / TMT-A | Aebi [18] | |
| Inhibition | Stroop-I | MNND Stroop Interference | Balzer, Berger, Caprez, Gonser, Gutbrod and Keller [19] | X Version A or B |
| Planning | ToL | MNND Planungstest (adapted version of the Tower of London) | Balzer, Berger, Caprez, Gonser, Gutbrod and Keller [19] | X Version A or B |
| Logical Reasoning | LPS-4 | Leistungsprüfsystem 50+: Subtest 4, Fluid Reasoning | Sturm, Willmes and Horn [20] | X A or B |
| **Attention & Working Memory** | | | | |
| Working Memory | DSback | Digit Span backwards (from Wechsler Adult Intelligence Scale (WAIS)) | Wechsler [21] | |
| Attention & Processing Speed | TMTA | CERAD Trail Making Test A | Aebi [18] | |
| | Stroop-R | MNND Stroop Word Reading | Balzer, Berger, Caprez, Gonser, Gutbrod and Keller [19] | X Version A or B |
| | Stroop-N | MNND Stroop Color Naming | Balzer, Berger, Caprez, Gonser, Gutbrod and Keller [19] | X Version A or B |
| **Memory** | | | | |
| Short-term memory | DSforw | Digit Span forwards (from Wechsler Adult Intelligence Scale (WAIS)) | Wechsler [21] | |
| | AVLGT-Learn | MNND Auditiv-Verbaler Lern- und Gedächtnistest AVLGT: Wordlist Learning | Balzer, Berger, Caprez, Gonser, Gutbrod and Keller [19] | X Version A or B |
| Verbal episodic long-term memory | AVLGT-Rec | MNND Auditiv-Verbaler Lern- und Gedächtnistest AVLGT: Wordlist Recall | Balzer, Berger, Caprez, Gonser, Gutbrod and Keller [19] | X Version A or B |
| Visuo-spatial episodic long-term memory | RTCFT-Rec | MNND Rey / Taylor Complex Figure Test (RTCFT): Figure Recall | Balzer, Berger, Caprez, Gonser, Gutbrod and Keller [19] | X Rey or Taylor |
| **Visuo-cognition** | | | | |
| Visuo-constructive abilities | RTCFT-Copy | MNND Rey / Taylor Complex Figure Test (RTCFT): Figure Copy | Balzer, Berger, Caprez, Gonser, Gutbrod and Keller [19] | X Rey or Taylor |
| Visuo-perceptive abilities | BJLO | Benton Judgment of Line Orientation | Benton, Hannay and Varney [22], Benton [23] | X Version V or H |
| **Language** | | | | |
| Naming | BNT | CERAD Boston Naming Test | Aebi [18] | |
| Semantic Vocabulary | Sim | Similarities (from Wechsler Adult Intelligence Scale (WAIS)) | von Aster and Neubauer [24] | |
| **Current vigilance state and inhibitory control** | | | | |
| Momentary Sleepiness | KSS | Karolinska Sleepiness Scale | Shahid, Wilkinson, Marcu and Shapiro [13], Akerstedt and Gillberg [14] | |
| Vigilance | SART | Sustained Attention to Response Task | Fronczek, Middelkoop, van Dijk and Lammers [15] | |
| Sustained attention | PVT | Psychomotor Vigilance Task | Thomann, Baumann, Landolt and Werth [16] | |
| Inhibitory Control | GNG | Go/NoGo Task | Fronczek, Middelkoop, van Dijk and Lammers [15] | |

*Notes.* CERAD, Consortium to Establish a Registry for Alzheimer's Disease; MNND, Materialien und Normwerte für die Neuropsychologische Diagnostik

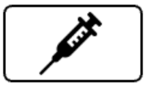

**Blood sampling.** At each study visit, i.e. three times in total, 20-30 ml of blood will be collected according to standard clinical practice in a fasting condition in the morning. Blood samples will be immediately centrifuged, aliquoted, and frozen and stored at -80°C until further analysis. The analysis will be carried out by investigators blinded to the study condition. Analyses will be performed using commercially available essays for measuring NfL (secondary outcome) and other biomarkers relevant in neurodegeneration (e.g. alpha-synuclein, tau, phospho-tau, Aß40, Aß42, synaptic and inflammatory markers; all exploratory outcomes). Due to rapid developments in the field, the final selection of exploratory blood-based biomarker outcomes and assays will be determined at the time of analysis.

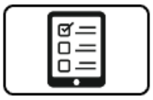

**Participant Reported Outcome Measures (PROMs).** The questionnaires assessing exploratory PROMs will be assessed during the clinical visits, and for some sleep-related PROMs (ESS, PDSS-2) additionally during in the intervention once per week, on a tablet. Table 2 lists the questionnaires and corresponding references.

*Table 2.* Questionnaires in PD-CogT-Sleep

| Domain | Abbreviation | Full questionnaire name | Reference |
|---|---|---|---|
| Subjective Cognitive Decline | Multi-SubCoDE | Multi-Domain Subjective Cognitive Decline Evaluation | Ophey, Röttgen, Doppler, Scharfenberg, Kufer, Farrher, Fink, Sommerauer and Kalbe [25] |
| Quality of Life | PDQ-39 | Parkinson’s Disease Questionnaire 39 | Peto, Jenkinson, Fitzpatrick and Greenhall [26],Berger, Broll, Winkelmann, Heberlein, Müller and Ries [27] |
| Fatigue | FSS | Fatigue Severity Scale | Krupp, LaRocca, Muir-Nash and Steinberg [28] |
| Depressive Symptoms & Anxiety | HADS | Hospital Anxiety and Depression Scale (HADS) | Zigmond and Snaith [29] |
| Sleep Disturbances | PDSS-2 | Parkinson’s Disease Sleep Scale 2 | Trenkwalder, Kohnen, Högl, Metta, Sixel-Döring, Frauscher, Hülsmann, Martinez-Martin and Chaudhuri [7] |
| Daytime Sleepiness | ESS | Epworth Sleepiness Scale | Johns [9] |
| Visual analogue scales | VAS | for daytime sleepiness, sleep quality, mood | e.g. used in Schreiner, Horlacher, Fattinger, Renerts, Brogli, Kämpf, Scandella, Poryazova, Valko, Ferster, Sassenburg, Jaramillo, Poian, Lustenberger, Karlen, Huber, Baumann and Maric [30] |

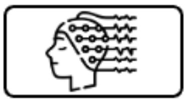

**Sleep high-density electroencephalography (hdEEG)**. All three clinical visits include a night of hdEEG recording. The hdEEG is conducted using a 128-lead hydrocell geodesic sensor net (HCGSN; EGI: Electrical Geodesics, Inc., Eugene, OR). From the hdEEG, we will derive local sleep EEG parameters, focused on SWA. Thereby, we can assess the effects of CogT on local SWA in the CogT+Sham group, and the effects of CogT+PTAS on local SWA in the CogT+PTAS group.

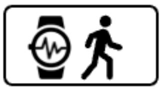

**Actigraphy.** Physical activity, sleep-wake patterns, and circadian rhythm will be measured with GENEActiv actigraphy devices from Activinsights over 2 weeks (14 days, 336 hours) starting at the first clinical visit and over 1 week prior to the second clinical visit (7 days, 168 hours). Measurements will be conducted with a sampling rate of 30 Hz with an accelerometer range of ± 8 g. The GENEactiv actigraphy device is CE-certified and has been validated as a measurement tool for classifying the intensity of physical activity and to examine sleep parameters.[31-33] The sleep-wake rhythm estimates from the actigraphy device will be used to confirm the self-reported sleep/wake times of the participants. We will furthermore assess parameters of physical activity in exploratory analyses. Data will be saved in the Continuous Wave Accelerometer (*.cwa) binary format containing the raw actigraphy data, metadata, and device configurations.

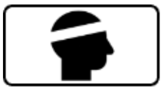

**Multi-night frontal sleep EEG.** We will derive multi-night parameters from sleep macro (i.e. sleep architecture, e.g. sleep stages and continuity) and microstructure (i.e. SWA) from the frontal EEG of the sleep headband (TOSOO Axora device), which is applied over multiple nights during interventions. In the CogT+Sham group, these parameters will be used to assess the effects of CogT on sleep. In the CogT+PTAS group, these parameters will be used to assess the effects of PTAS on sleep in participants applying CogT.

**Sleep and application diary.** A sleep diary, and application diary will be kept according to protocols previously conducted at our study site.[30]. Every evening during the intervention period, participants will complete a short checklist ensuring proper application of the sleep device. Every morning during the intervention period, participants will complete a short digital questionnaire about the nightly experience (visual analogue scale for sleep quality, KSS, application of the sleep device and potential problems, e.g., technical errors). Additionally, participants will be asked to report PD-related sleep disturbances (PDSS-2[7]) and daytime sleepiness (ESS[9]) on a weekly basis.
